# Global DNA hypomethylation in epithelial ovarian cancer: passive demethylation and association with genomic instability

**DOI:** 10.1101/2020.01.22.20018374

**Authors:** Wa Zhang, David Klinkebiel, Carter J Barger, Sanjit Pandey, Chittibabu Guda, Austin Miller, Stacey N. Akers, Kunle Odunsi, Adam R. Karpf

## Abstract

A hallmark of human cancer is global DNA hypomethylation (GDHO), but the mechanisms accounting for this defect and its pathological consequences have not been defined in human epithelial ovarian cancer (EOC). In EOC, GDHO was associated with advanced disease and reduced overall and disease-free survival. GDHO(+) EOC was enriched for a proliferative gene expression signature, including CCNE1 and FOXM1 overexpression. DNA hypomethylation preferentially occurred within genomic blocks (hypomethylated blocks) overlapping late-replicating, lamina-associated domains, PRC2 binding, and H3K27me3. Increased proliferation coupled with hypomethylated block formation at late replicating regions suggested passive hypomethylation, which was further supported by the observation that cytosine DNA methyltransferases (*DNMTs)* and *UHRF1* showed significantly reduced expression in GDHO(+) EOC, after normalization to proliferation markers. Importantly, GDHO(+) EOC showed elevated chromosomal instability (CIN), and copy number alterations (CNA) were enriched at hypomethylated blocks. Together, these findings implicate a passive demethylation mechanism for GDHO that promotes genomic instability and poor prognosis in EOC.

---

Altered DNA methylation, a fundamental characteristic of human cancer, includes gains and losses of methylation [1,2]. DNA hypermethylation leads to tumor suppressor gene silencing and occurs frequently at genomic regions occupied by polycomb group proteins in ES cells [1,3]. In contrast, DNA hypomethylation is referred to as “global,” as 5-methyl-deoxycytidine (5mdC) levels are often reduced in cancer [4-6]. In agreement, DNA hypomethylation occurs at repetitive elements (RE), including the interspersed retrotransposon *LINE-1* [7], which accounts for ∼17% of the genome. Global DNA hypomethylation (GDHO) is also associated with hypomethylation and activation of cancer-testis or cancer-germline (CG) genes [8-13]. More recently, epigenomic approaches have revealed that global DNA hypomethylation is not random or driven solely by changes at RE, but rather is localized at large genomic regions called hypomethylated blocks [14-16]. Hypomethylated blocks overlap lamina-associated domains (LADs) and, interestingly, appear to contain epigenetically silenced genes, as well as genomic regions showing high gene expression variability [15,17,18].

GDHO, which is commonly measured using RE methylation markers such as *LINE-1*, is associated with poor clinical outcome, but the reasons for this association have not been established [19]. A number of plausible mechanisms could account for this link. First, GDHO may promote chromosomal instability (CIN), as genetically induced DNA hypomethylation in mouse tumor models and human cancer cell lines leads to aneuploidy, chromosomal translocations, and copy number alterations (CNA) [20-22]. Supporting this model, DNA hypomethylation and genomic alterations are linked in human cancer [23-28]. Second, aberrant gene expression, including oncogene activation, RE expression, or CG antigen gene activation may promote oncogenic phenotypes and/or disease progression [2,29-32]. Third, GDHO and the associated hypomethylated block formation could promote gene expression variability and provide a selective growth advantage, e.g. chemotherapy resistance or accelerated disease progression [33].

In addition to identifying the targets and consequences of GDHO, it is of high interest to understand its origin. At least two general types of mechanisms may underlie GDHO. First, “active” hypomethylation, caused by a specific molecular alteration that disrupts DNA methylation or enhances DNA demethylation, could be involved. Potential related mechanisms could include, for example, mutations in *DNMTs* or *Ten-eleven translocation methylcytosine dioxygenase* (*TET*) genes. Second, “passive” hypomethylation is predicated on the fact that DNA methylation is a post-replicative DNA modification that could become unlinked from DNA replication. In the passive model, hypomethylation is proposed to indirectly result from cellular transformation and the accompanied increase in cell proliferation [34]. A recent important study demonstrated a link between DNA hypomethylation, late replication, and mitotic cell division in many human cancers [28]; however, little information is currently available for EOC.

RNA sequencing (RNA-seq) is commonly used to study promoter variation and splice variants, which are difficult to measure using microarrays [35]. It is less appreciated that RNA-seq can also measure RE-derived transcripts, which is excluded from microarray designs and are traditionally omitted from studies of the cancer transcriptome. Notably, total RNA-seq revealed frequent and widespread expression of RE in pancreatic cancer and other tumors [36]. The mechanisms accounting for RE expression in cancer are unknown, but could potentially include epigenetic activation by DNA hypomethylation.

Epithelial ovarian cancer (EOC), and specifically the high-grade serous subtype, HGSOC, is the most lethal gynecologic malignancy, due to a lack of diagnostic approaches coupled with poor long-term survival for patients with late stage disease [37]. EOC is characterized by widespread CNA, *TP53* mutations, defects in homologous recombination (including BRCA genes), retinoblastoma protein (RB) pathway dysregulation, and FOXM1 activation [38]. In addition to genetic changes, EOC shows altered DNA methylation, including both hyper- and hypomethylation[39]. More specifically, GDHO, including *LINE-1* hypomethylation is a common phenotype observed in EOC [9,40,41]. In addition, *LINE-1* hypomethylation and expression was recently shown to be a common event in ovarian cancer precursor lesions, known as serous tubal intraepithelial carcinoma (STICs) [42]. Here, we studied GDHO in EOC, including its clinic-pathological context, molecular underpinnings, and relationship to CIN.

## Results

### *LINE-1* hypomethylation is associated with disease progression and reduced survival in EOC

Previously, we reported that *LINE-1* methylation is an appropriate biomarker for global DNA methylation in EOC, and that *LINE-1* is hypomethylated in EOC as compared to bulk normal ovary (NO), ovarian surface epithelia (OSE), or fallopian tube epithelia (FTE) tissues[9,43]. Here, we investigated the relationship between *LINE-1* methylation and EOC clinic-pathology. Notably, *LINE-1* hypomethylation was associated with increased clinical stage and histopathological grade, and correlated with reduced overall and disease-free survival (Fig. 1a-d). Furthermore, the association with disease free survival remained significant in a proportional hazards model after adjustment for age (HR P=0.03).

**Figure 1.**
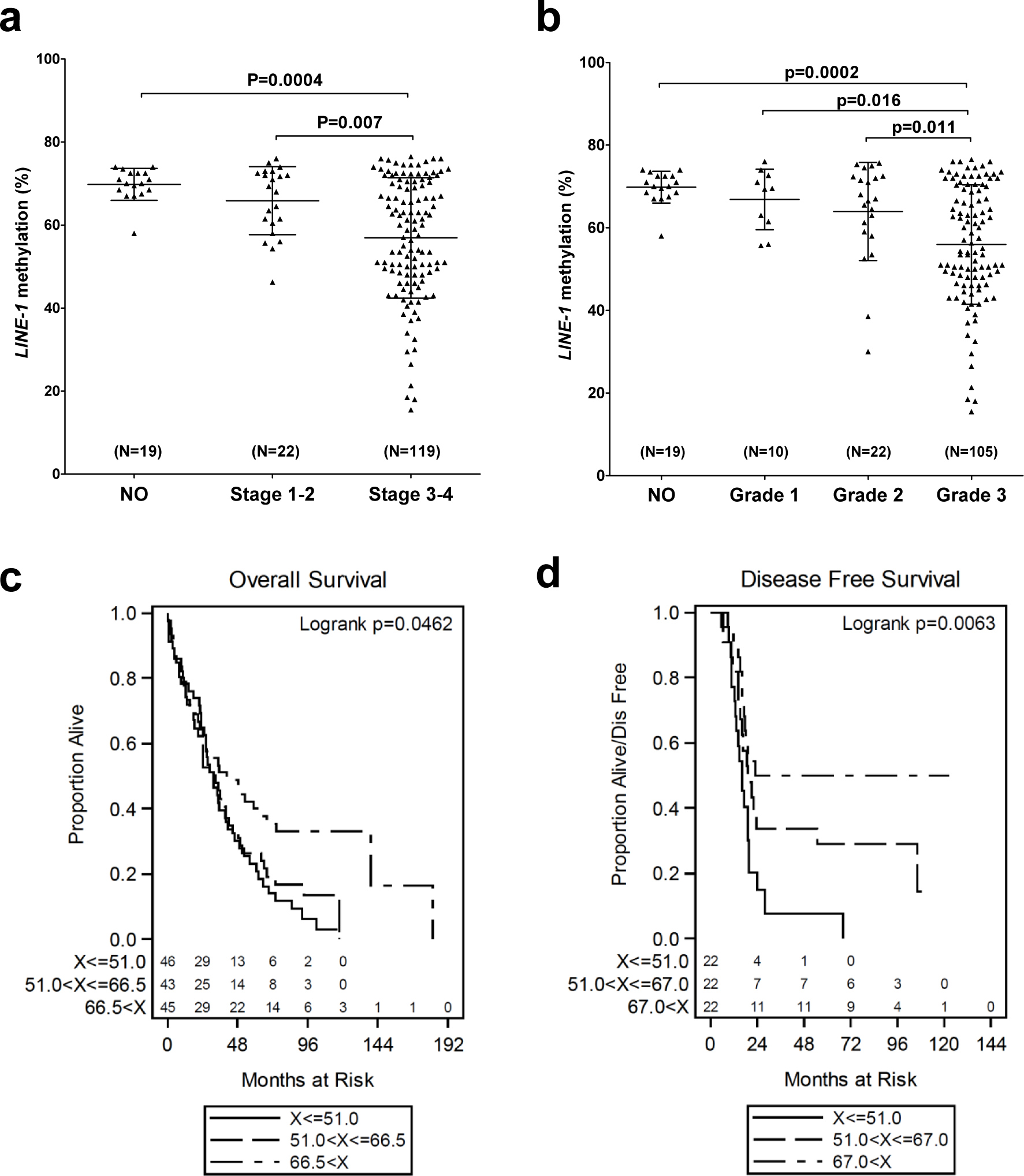
*LINE-1* hypomethylation is associated with EOC disease progression and reduced survival. *LINE-1* methylation was determined by sodium bisulfite pyrosequencing. **(a)** *LINE-1* methylation vs. clinical stage. (**b)** *LINE-1* methylation vs. pathological grade. For **a** and **b**, mean +/- SD is plotted, and Mann Whitney p-values are indicated. **(c**,**d)** Kaplan-Meier survival analyses and log rank test p-values of EOC patients separated based on tumor *LINE-1* methylation values. **(c)** *LINE-1* methylation vs. overall survival. Patients were separated into three groups based on *LINE-1* methylation values: low (<51.0%), middle (51.0%-66.5%), and high (>66.5%). A key for the three groups, and the number of patients in each, is shown. **(d)** *LINE-1* methylation vs. disease-free survival. Patients were separated into three groups based on *LINE-1* methylation values: low (<51.0%), middle (51.0%-67.0%), and high (>67.0%). A key for the three groups, and the number of patients in each, is shown.

### Global gene expression patterns are distinct in GDHO(+) EOC

We used gene expression microarrays to profile: i) EOC samples showing significant *LINE-1* hypomethylation [GDHO(+) EOC; N=20], ii) EOC samples with *LINE-1* methylation levels similar to NO [GDHO(–) EOC; N=20], and iii) NO; N=3 (Fig. 2a). For expression studies, we used NO as a control because of difficulty in obtaining high quality RNA from primary OSE and FTE tissues. Hierarchical clustering of differentially expressed genes (DEG) revealed distinct patterns of gene expression in GDHO(+) and GDHO(–) EOC (Fig. 2b). Using a cutoff of P<0.01, 1696 Affymetrix probe sets (genes) were differentially expressed in the two EOC groups, with 958 (56%) of these upregulated in GDHO(+) EOC (Fig. 2b; Supplementary Table 1). Clustering of DEG using a false discovery rate (FDR) cutoff of 0.1 resulted in a similar separation of GDHO(+) from GDHO(−) samples (Supplementary Fig. 1a). Because GDHO is associated with disease progression (Fig. 1), we conducted sub-group analysis using disease-matched tumors, in which we compared age-matched, stage 3/4, grade 3, serous EOC (i.e. HGSOC) samples from each group [GDHO(+) N=13; GDHO(−) N=9]. Importantly, disease matched GDHO(+) and GDHO(–) EOC gene expression patterns remained distinct (and provided improved group separation) in this comparison (Fig. 2c). At P<0.01, 752 Affymetrix probe sets (genes) were differentially expressed, with 357 (47%) upregulated in GDHO(+) EOC (Fig. 2C; Supplementary Table 1). Clustering of DEG using a false discovery rate (FDR) cutoff of 0.1 resulted in similar separation of disease-matched GDHO(+) vs. GDHO(−) EOC (Supplementary Fig. 1b). A list of all DEG identified in both comparisons [GDHO(+) vs. GDHO(−): overall and disease-matched samples] is provided in Supplementary Table 2, and the microarray raw intensity files have been deposited in GEO (see *Methods*).

**Fig 2.**
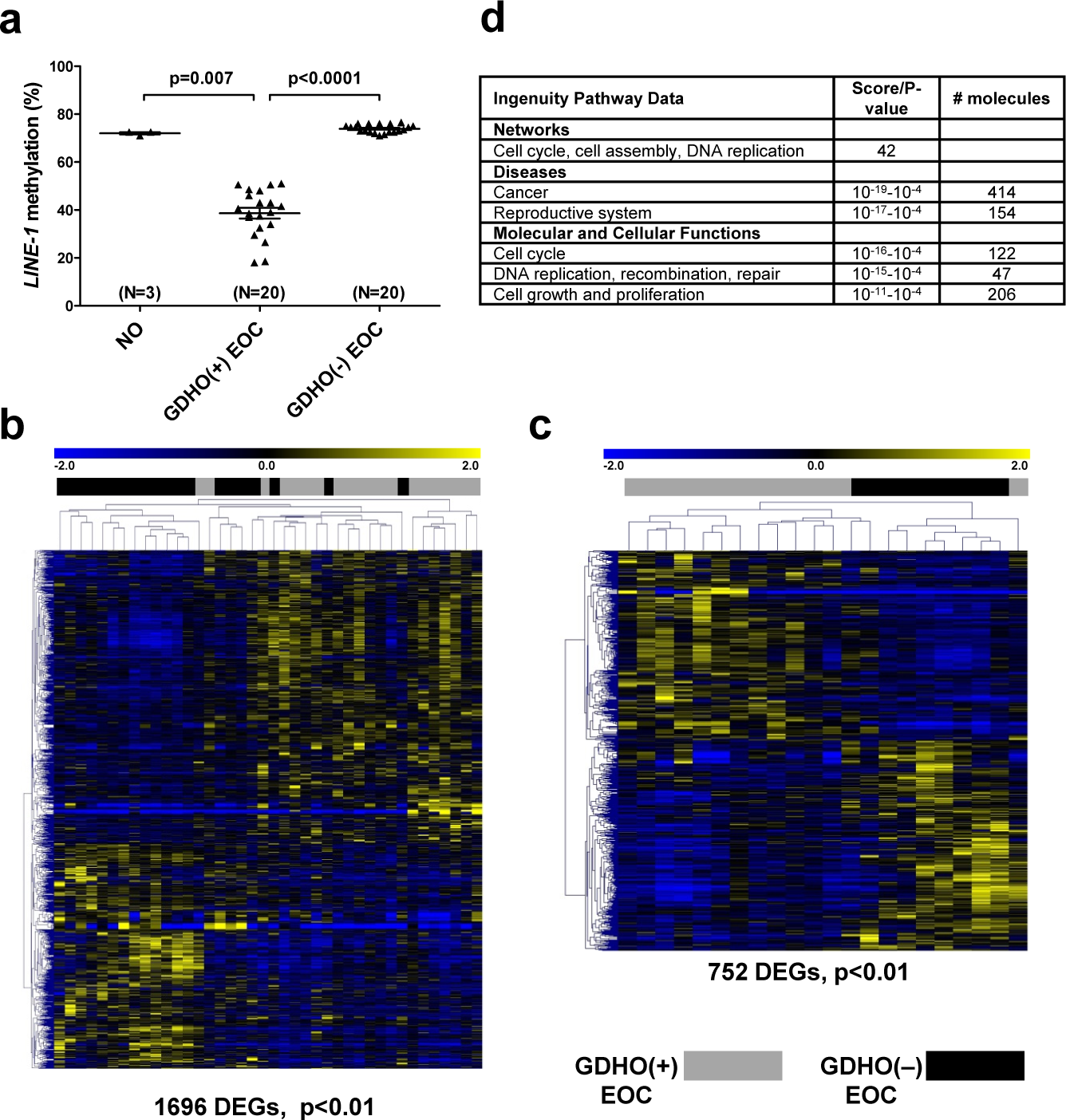
Distinct patterns of gene expression in GDHO(+) EOC. **(a)** *LINE-1* methylation in sample groups used for Affymetrix gene expression analyses: bulk normal ovary (NO), GDHO(+) EOC (*LINE-1* hypomethylated), GDHO(–) EOC (*LINE-1* hypermethylated). **(b)** Hierarchical clustering heat map of genes differentially expressed (p<0.01) between GDHO(+) vs. GDHO(–) EOC (all samples). **(c)** Hierarchical clustering heat map of genes differentially expressed between disease-matched (age-matched, stage 3/4, grade 3, serous EOC) GDHO(+) vs. GDHO(–) EOC. The number of genes showing significant differential expression in each comparison is indicated. Sample identities are indicated at top (see key). **(d)** Selected IPA data for the GDHO(+) vs. GDHO(–) EOC microarray comparison.

### Proliferative expression signature and FOXM1 activation in GDHO(+) EOC

We used Ingenuity Pathway Analysis (IPA) of microarray expression data to identify cellular pathways involved in GDHO. We observed significant alterations in both cancer and reproductive disease pathways (Fig. 2d; Supplementary Table 3). In addition, functional pathways related to cell growth, proliferation, and division were altered; together these data suggested that increased proliferation is a key characteristic of GDHO(+) EOC (Fig 2d; Supplementary Table 3). To test this, we measured the expression of the canonical cell proliferation marker *MKI67[44]* in GDHO(+) and GDHO(–) EOC. Both microarray and RT-qPCR analysis demonstrated significantly elevated *MKI67* expression in GDHO(+) EOC, as well as a significant negative association with *LINE-1* methylation (Supplementary Fig. 2). In addition, two key drivers of EOC proliferation, *CCNE1* and *FOXM1*, were markedly upregulated in GDHO(+) EOC (Fig. 3a,b). An association of *CCNE1* amplification with DNA hypomethylation has been noted previously in stomach cancer [45]. Based on the emerging role of the FOXM1 pathway in EOC and a report that FOXM1 may be linked to changes in DNA methylation[38,46], we tested the association between *FOXM1* expression and *LINE-1* methylation using an expanded set of EOC samples. Notably, the data revealed a strong inverse association (Fig. 3c). In addition to the mRNA, expression of the FOXM1 protein and also FOXM1 target genes involved in cell cycle progression, including *PLK1, AURKB, BIRC5*, and *CCNB1*[38], were significantly upregulated in GDHO(+) EOC, consistent with functional activation of FOXM1 (Fig 3d; Supplementary Fig. 3). Furthermore, gene set enrichment analysis (GSEA) indicated a significant enrichment of the FOXM1 transcription factor network (p <0.01, FDR <0.25) and G2/M checkpoint genes (p < 0.05, FDR <0.25) in GDHO(+) EOC. Together, these data implicate increased FOXM1 expression as a prominent feature of GDHO in EOC. Activation of *CCNE1* and *FOXM1* in GDHO(+) EOC could potentially result from direct hypomethylation of their promoter regions, resulting from a passenger effect in GDHO(+) tumors. However, bisulfite sequencing analyses indicated that the *CCNE1* and *FOXM1* promoters were constitutively hypomethylated in NO, GDHO(–) and GDHO(+) EOC (Supplementary Fig. 4), suggesting that increased proliferation may promote GDHO, rather than *vice versa*.

**Fig 3.**
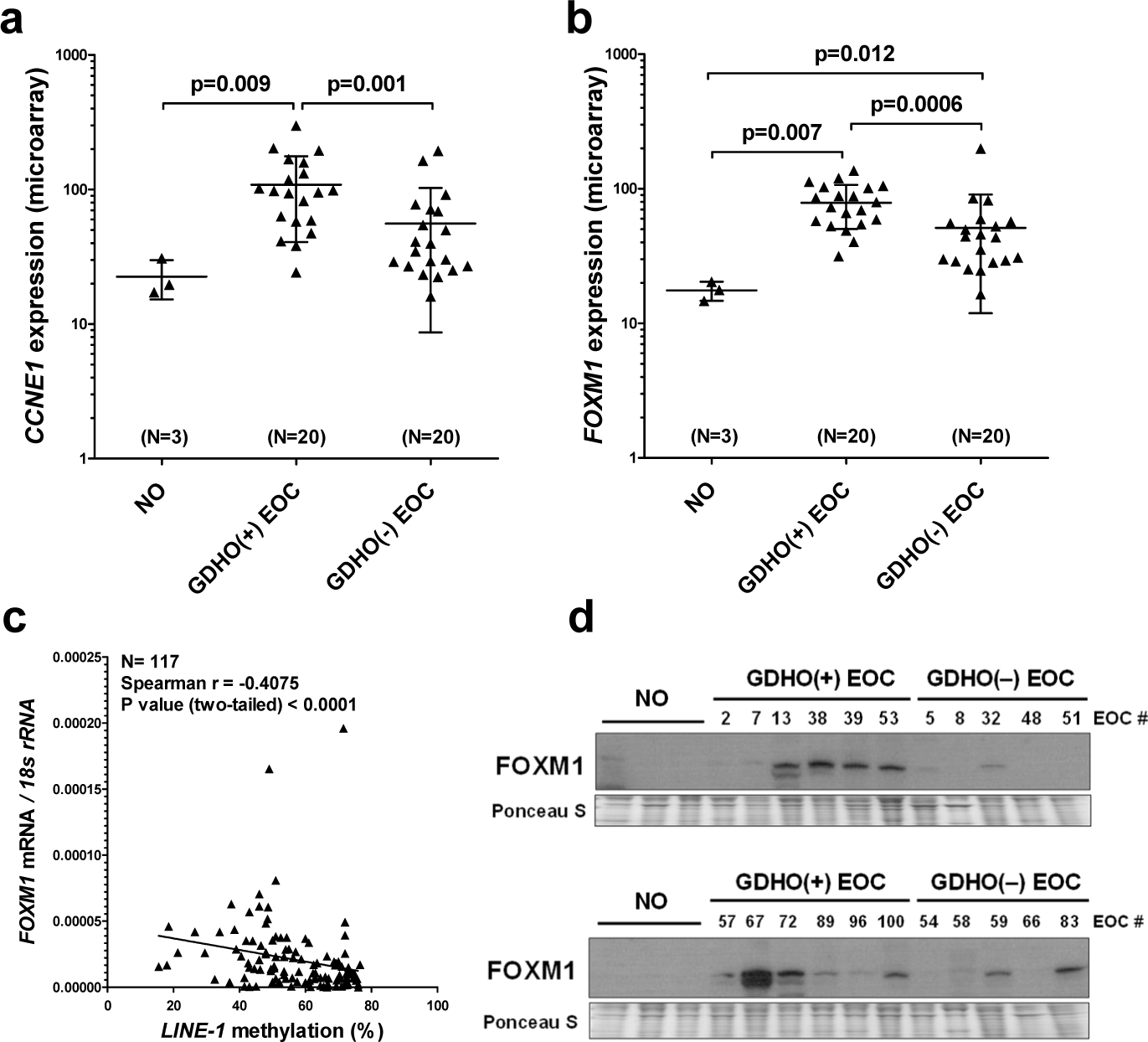
*CCNE1* and FOXM1 overexpression in GDHO(+) EOC. **(a)** *CCNE1* expression in NO, GDHO(+) EOC, and GDHO(–) EOC, as determined by Affymetrix microarray. **(b)** *FOXM1* expression in NO, GDHO(+) EOC, and GDHO(–) EOC, as determined by Affymetrix microarray. Means +/− SD are plotted, and Mann Whitney p-values are indicated. **(c)** *FOXM1* mRNA expression vs. *LINE-1* methylation in EOC. *FOXM1* expression was measured by RT-qPCR while *LINE-1* methylation was measured by pyrosequencing. Spearman test results and p-value are shown. **(d)** Western blot analysis of FOXM1 protein expression in NO, GDHO(+) EOC, and GDHO(–) EOC. The two blots are comprised of different sets of samples. Ponceau S staining is shown as a loading control.

### Widespread activation of cancer germline (CG) genes and altered expression of epigenetic regulators and histone genes in GDHO(+) EOC

Expression of CG genes in association with GDHO in cancer has been reported, but previous studies have only studied one or a limited number of genes[8-10]. Our current data set allowed for a comprehensive and global examination of this association. Almost one-fifth of annotated CG genes were differentially expressed in GDHO(+) vs. GDHO(−) EOC, with each showing upregulation (Fig. 4; Supplementary Table 4). In agreement, GSEA analysis of CG genes showed significant enrichment in GDHO(+) EOC (p<0.01, FDR<0.25). Over half of the CG genes activated in GDHO(+) EOC were also induced in the disease-matched comparison, including both X-chromosome and autosomal genes (Fig. 4; Supplementary Table 4). The differentially expressed CG genes (DE-CG genes) included cancer vaccine targets and genes with oncogenic function, including *MAGEA* genes, *NY-ESO-1, XAGE-1*, and *PRAME* (Supplementary Table 4). To test whether these genes are directly regulated by DNA methylation, we used Affymetrix microarrays to measure their expression in EOC cell models before and after decitabine (DAC) treatment. Notably, the majority of the DE CG genes were upregulated following decitabine treatment (Fig 4; Supplementary Table 4). This observation, coupled with extensive prior data regarding CG gene hypomethylation in cancer[10], strongly suggests that many of the DE CG genes are directly regulated by DNA methylation.

**Fig 4.**
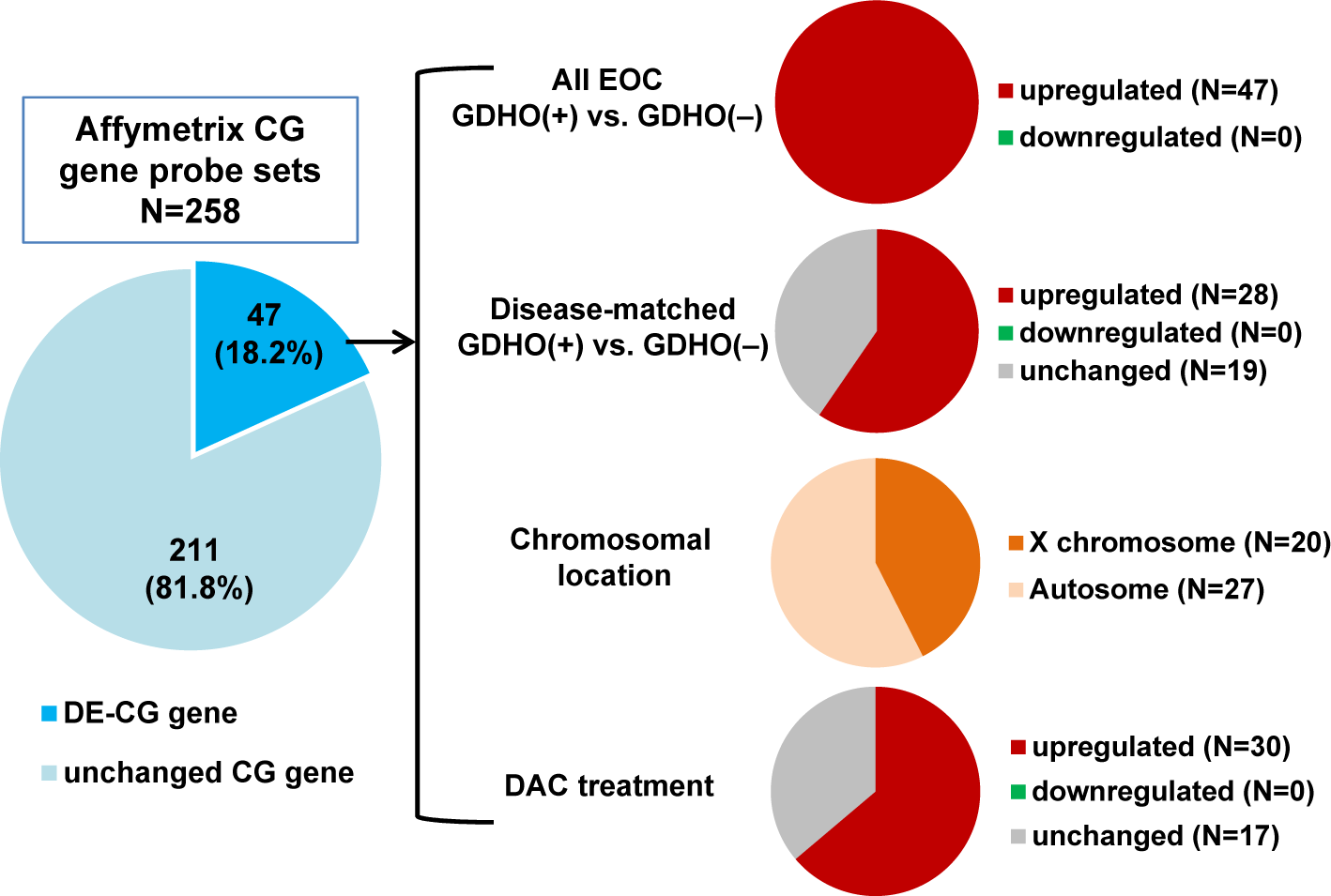
Differential expression of CG antigen genes in GDHO(+) EOC and regulation by DNA methylation. CG antigen gene list was obtained from the CT gene database on March 19, 2015: http://www.cta.lncc.br/. **Left:** the proportion of CG genes (i.e., the corresponding Affymetrix probes) differentially expressed between GDHO(+) and GDHO(–) EOC (p<0.01) as determined by Affymetrix microarray. **Right:** Direction of expression change for the DEGs in all samples or disease-matched samples, their chromosomal localization, and the effect of decitabine (DAC) treatment on their expression in EOC cell lines, as determined by Affymetrix microarray.

Two additional gene groups of note had altered expression in GDHO(+) EOC. First, genes with known roles in epigenetic regulation were altered. These genes included *EHMT2*/*G9a, ATAD2*, and *HDAC1*, which have reported oncogenic activity in EOC[47-51] (Supplementary Table 5). Increased expression of *ATAD2* in GDHO(+) tumors could be due to promoter hypomethylation[52]. Interestingly, there was significantly increased expression both of epigenetic regulators involved in gene activation (e.g. *JMJD2a, ATAD2, ASF1b*), as well as gene repression (e.g. *G9a, HDAC1, LSD1*) in GDHO(+) EOC (Supplementary Table 5). These changes may reinforce the epigenetic impact of GDHO, or alternatively could become activated as a feedback inhibitory response to GDHO. In addition to direct epigenetic regulators, we noted that approximately half of all histone genes were upregulated in GDHO(+) EOC as compared to GDHO(−) EOC (Supplementary Table 6). In part, this could reflect the increased proliferation signature observed in these tumors; in addition, direct hypomethylation of histone genes in EOC has recently been reported[53]. Bisulfite sequencing analysis of select histone genes indicated that some are constitutively hypomethylated in NO and EOC, while others are hypomethylated in GDHO(+) EOC, as compared to GDHO(−) EOC (Supplementary Fig. 5).

### DNA Methylome analysis of GDHO(+) EOC

For initial analysis of the DNA methylation landscape of GDHO(+) EOC, we used Illumina Infinium 450K bead arrays (450K)[54]. In addition to profiling both groups of EOC, we analyzed normal epithelia (NE; OSE + FTE average) as a control. Both unsupervised hierarchical clustering and principal component analyses (PCA) of 450K data revealed that GDHO(+) and GDHO(–) EOC have distinct DNA methylomes, and that each are distinct from NE (Fig. 5a,b). As a complementary approach, and to define the methylome in greater depth, we performed methylome sequencing (Methyl-seq) using a solution hybridization selection method[55] (Agilent SureSelect). 450K and Methyl-seq data were highly concordant (Pearson r = 0.97; N=347,357 CpG sites), and hierarchical clustering and PCA analyses of Methyl-seq confirmed that GDHO(+) EOC, GDHO(–) EOC, and NE have distinct methylomes (Fig. 5c,d). As expected, DNA methylation overall was reduced in GDHO(+) EOC, as determined by either DNA methylome method (Fig. 5 e,f).

**Fig 5.**
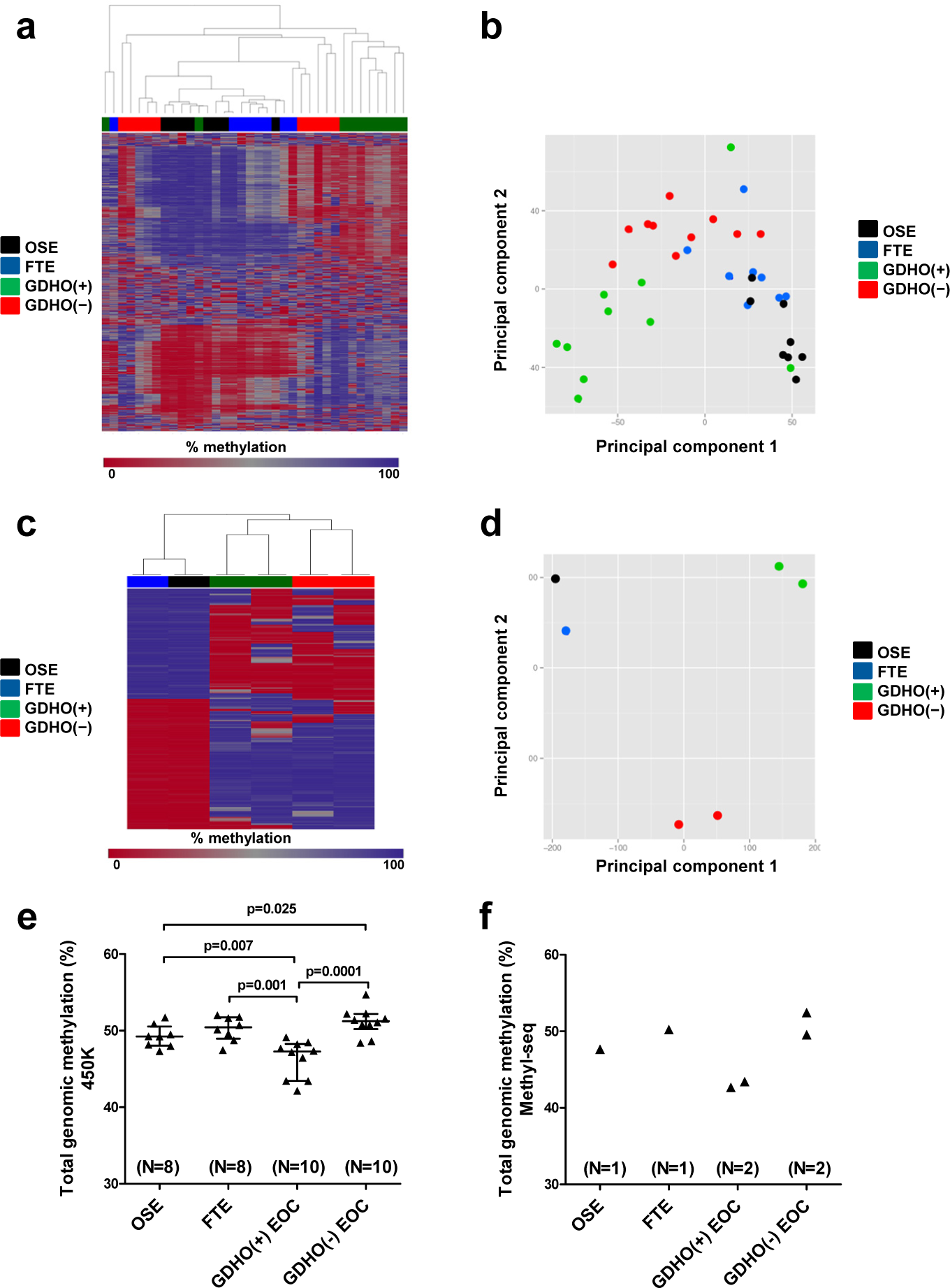
DNA methylome analysis of GDHO(+) EOC. **(a)** Dendogram showing relatedness of OSE, FTE, GDHO(+) EOC, and GDHO(–) EOC methylomes, using Illumina 450K beadarray analysis. Data for the 1000 most variable CpG sites is shown. **(b)** Principal component analysis of sample groups using all 450K data. **(c)** Dendogram showing relatedness of OSE, FTE, GDHO(+) EOC, and GDHO(–) EOC methylomes, using Agilent Sure Select Methyl-seq. The 1000 most variable CpG sites is shown. **(d)** Principal component analysis of sample groups using all Methyl-seq data. **(e)** Total genomic methylation of OSE, FTE and EOC sample groups as determined by 450K. Median and interquartile ranges are plotted, and the unpaired two-tailed t-test p-values are shown. **(f)** Total genomic methylation levels of OSE, FTE and EOC sample groups as determined by Methyl-seq.

Both 450K and Methyl-seq indicated that a large number of promoters were significantly differentially methylated in the two EOC groups, with a vast majority, regardless of CpG island context, showing hypomethylation in GDHO(+) EOC (Supplementary Fig. 6). In contrast, promoter hypermethylation and hypomethylation was almost evenly split when comparing EOC to NE, and, strikingly, CpG island promoters were mostly hypermethylated while non-CpG island promoters were mostly hypomethylated in EOC (Supplementary Fig. 6). We also defined differentially methylated regions (DMR) globally and found that the vast majority of DMR were hypomethylated in GDHO(+) as compared to GDHO(−) EOC (Supplementary Fig. 7). The majority of DMR were also hypomethylated in EOC as compared to the NE control (Supplementary Fig. 7). As methylation at different genomic locations has distinct consequences, we calculated differentially methylated CpGs (DMC) independently for genes, CpG islands, CpG shores, CpG shelves, and CpG open seas, as described previously[54]. Most DMC were hypomethylated in GDHO(+) as compared to GDHO(–) EOC, regardless of genomic context (Supplementary Fig. 7 c,d), while hypermethylation and hypomethylation was more evenly split for EOC vs. NE (Supplementary Fig. 7 e,f). In the latter comparison, CpG rich regions of the genome favored hypermethylation in EOC, while CpG poor regions (i.e. open seas) favored hypomethylation in EOC.

### Hypomethylated genomic blocks in GDHO(+) EOC

To investigate whether hypomethylated blocks[14,15] are present in EOC, and to determine their relationship to GDHO, we first visually inspected Methyl-seq data using the *UCSC Genome Browser*. Examples of the resulting data are shown for chromosome 11 (Fig. 6a,b). These observational data suggested that hypomethylated blocks were present and enriched in GDHO(+) EOC. To formally test this, we used a quantitative approach to determine the number and size of hypomethylated blocks in the different EOC groups, using NE as the control (see *Methods*). While both 450K and Methyl-seq were capable of detecting and quantifying hypomethylated blocks, we only present Methyl-seq data here, due to its significantly greater genomic coverage. We observed a large enrichment in both the number and size of hypomethylated blocks in GDHO(+) EOC as compared with GDHO(–) EOC, with approximately 14% of the genome residing in hypomethylated blocks in the former (Supplementary Table 7). These data indicate that hypomethylated block formation is heterogeneous in EOC, and may be closely linked to more common measures of GDHO, such as *LINE-1* hypomethylation. Based on previous findings in other cancer types, we next quantified the overlap between EOC hypomethylated blocks, lamina-associated domains (LADs), and late-replicating genomic regions. We observed significant enrichment of hypomethylated blocks at LADs, and a similar enrichment at late-replicating regions (Fig 6b; Table 1). Genes and CpG islands were also enriched in hypomethylated blocks, but this likely reflects the bias of our Methyl-seq approach to coverage of these regions (Table 1; Supplementary Table 8). We also analyzed hypomethylated blocks for overlap with specific transcription factor (TF) binding sites, histone modifications, and RE, using ENCODE and the UCSC genome browser database. Among TF binding sites showing strong enrichment in EOC hypomethylated blocks were the polycomb repressor complex 2 (PRC2) components EZH2 and SUZ12, and the genomic insulator CTCF (Table 1; Supplementary Table 9). Hypomethylated blocks were also highly enriched for H3K27me3, the modification recognized by the PRC2 complex (Table 1; Supplementary Table 10). These data suggest an association between DNA hypomethylation and repressive chromatin formation in EOC, as was reported previously in a breast cancer cell line[17]. Notably, hypomethylated blocks have been proposed to facilitate variable gene expression, which may provide a selective growth advantage during tumorigenesis[16]. In agreement, we found that genes with high expression variability were enriched in the EOC hypomethylated blocks (Fig 6c).

**Table 1.** Correlation of EOC hypomethylated blocks with genomic features^1, 2^.

| Genomic Region | # of regions | Size of regions (bp) | % of genome | % regions in hypo blocks | Correlation | Jaccard Test P-value<sup>3,4</sup> |
| --- | --- | --- | --- | --- | --- | --- |
| EOC Hypo Blocks | 2,208 | 8.81E+08 | 29 | 100 | N/A | N/A |
| LADs | 1,231 | 1.13E+09 | 37 | 36 | Direct | <0.01 |
| Early Replicating | 910 | 7.10E+08 | 23 | 25 | Indirect | <0.01 |
| Late Replicating | 1,155 | 5.81E+08 | 19 | 36 | Direct | <0.01 |
| CpG islands | 27,537 | 2.10E+07 | 1 | 32 | Direct | <0.01 |
| Genes | 28,489 | 1.41E+09 | 46 | 32 | Direct | <0.01 |
|  | # binding sites | # binding sites in hypo blocks | % binding sites in hypo blocks | Fold Enrichment | Correlation | Projection Test P-value |
| EZH2 Sites | 14,818 | 6,103 | 41 | 1.41 | Direct | <0.01 |
| SUZ12 Sites | 5,772 | 2,497 | 43 | 1.48 | Direct | <0.01 |
| CTCF Sites | 162,209 | 52,216 | 32 | 1.10 | Direct | <0.01 |
|  | # peaks | # peaks in hypo blocks | % peaks in hypo blocks | Fold Enrichment | Correlation | Jaccard Test P-value |
| H3K4me3 | 33,116 | 8,655 | 26 | 0.90 | Indirect | <0.01 |
| H3K9me3 | 49,328 | 18,264 | 37 | 1.28 | Direct | <0.01 |
| H3K27ac | 67,989 | 17,935 | 26 | 0.91 | Indirect | <0.01 |
| H3K27me3 | 40,126 | 15,428 | 38 | 1.32 | Direct | <0.01 |
| H3K36me3 | 33,342 | 6,303 | 19 | 0.65 | Indirect | <0.01 |
|  | # of repeats | # repeats in hypo blocks | % repeats in hypo blocks | Fold Change | Correlation | Jaccard Test P-value |
| *LINE-1* | 951,780 | 264,352 | 28 | 0.96 | Direct | <0.01 |
| *Alu/SINE* | 1,194,734 | 292,495 | 24 | 0.84 | Indirect | <0.01 |
| *Satellites* | 6,775 | 1,052 | 16 | 0.64 | Indirect | <0.01 |

**1** Hypo blocks were determined from Methyl-seq comparison of NE (OSE + FTE average) to GDHO(+) EOC
**2** Genomic features were retrieved from public databases, as described in *Methods*
**3** Statistical tests were performed using GenometriCorr (Favorov et. al, *PLoS Computational Biology*, 2012), with hypo block coordinates used as the reference value.
**4** All tests, other than CpG islands, gave the same result when using hypo blocks as query and the genomic features as the reference value. In contrast, the association with CpG islands was not significant.

**Fig 6.**
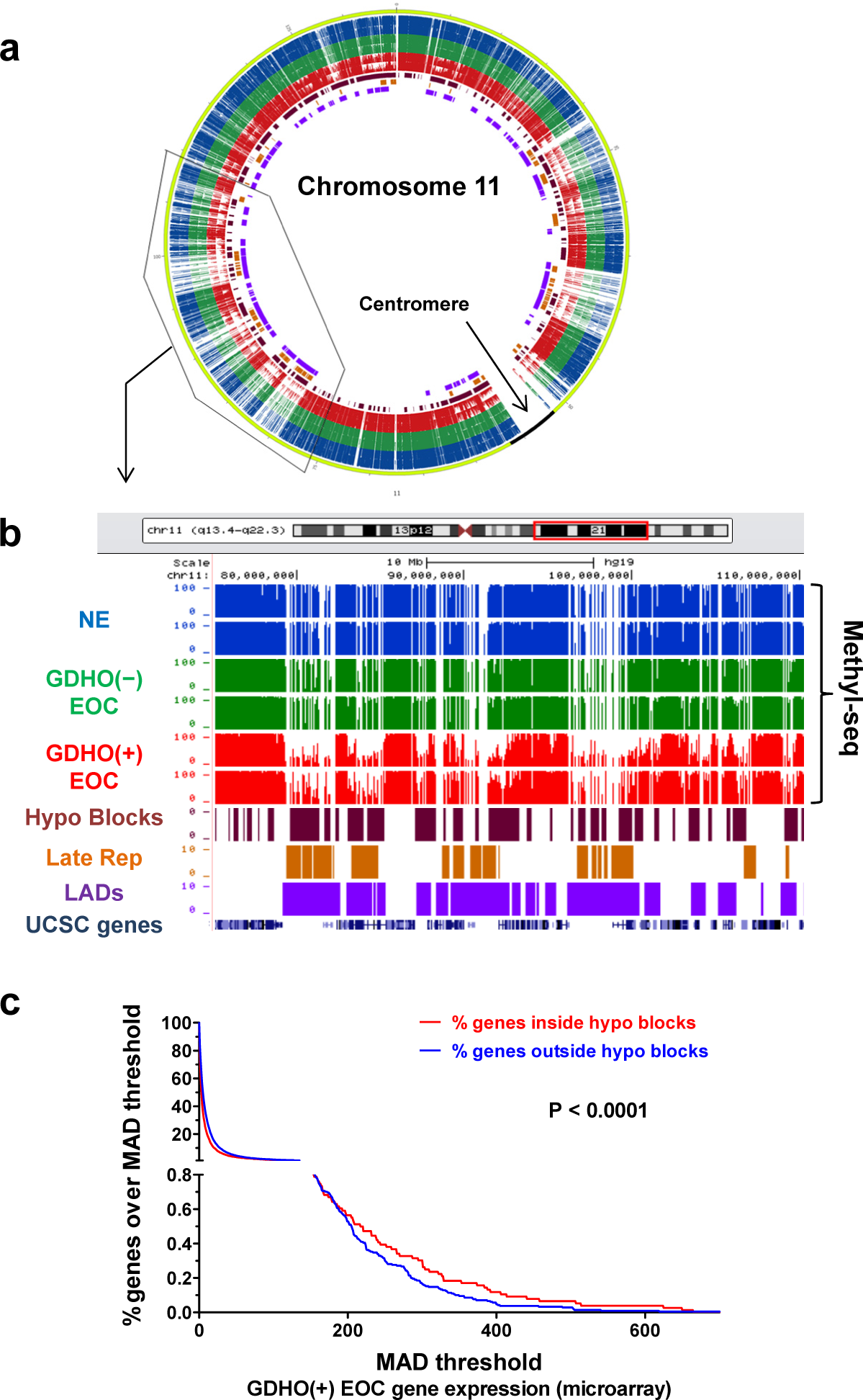
Hypomethylated genomic blocks (hypomethylated blocks) in GDHO(+) EOC. **(a)** Circos plot (http://circos.ca/) of Methyl-seq data of chromosome 11 for OSE, FTE, GDHO(+) EOC, and GDHO(–) EOC. Methyl-seq data for each sample group, hypomethylated blocks, late replicating regions, and LADs are indicated (see panel **b** for key). The enclosed region of the circos plot is enlarged in panel **b. (b)** Methyl-seq data for a selected region of chromosome 11 for OSE, FTE, GDHO(+) EOC, and GDHO(–) EOC. Calculated hypomethylated block regions are indicated at bottom, along with UCSC genome browser view of lamina-associated domains (LADs), late-replicating (Late rep) regions and gene positions. The Methyl-seq data is plotted on a scale from 0-100% methylation. **(c)** The relationship between hypomethylated blocks and gene expression variability in GDHO(+) EOC. Gene expression variability at hypomethylated blocks and non-block regions was determined from Affymetrix microarray data, by calculating the median average deviation (MAD) for individual gene expression values amongst the 20 GDHO(+) EOC samples. Hypomethylated blocks were calculated from Methyl-seq data by comparing GDHO(+) EOC to normal epithelia (NE; FTE + OSE average value). The 2-tailed paired t-test p-value is shown. The results indicate enrichment of hypervariable gene expression inside of hypomethylated blocks.

### Maintenance methylation components have reduced expression relative to proliferation markers in GDHO(+) EOC

The proliferative gene expression signature found in GDHO(+) EOC, coupled with the presence of hypomethylated blocks that overlap late-replicating regions, suggests a passive DNA hypomethylation mechanism. Paradoxically, however, elevated *DNMT* expression (relative to normal tissues) has been commonly observed in cancer. To address this discrepancy, we first analyzed *DNMT* expression in our samples using standard Robust Multichip Average (RMA) Affymetrix normalization. As expected, *DNMT1* and *DNMT3B* were upregulated in EOC compared to NO, while *DNMT3A* and *DNMT3L* were expressed at similar levels (Supplementary Fig. 8). However, it is important to note that maintenance DNA methylation is restricted to S phase, where *DNMT* expression is also increased[56]. Therefore, we hypothesized that *DNMT* expression normalized to cell proliferation status is a more relevant measure of maintenance methylation capacity in tumors. Remarkably, after normalization to the canonical cancer cell proliferation marker *MKI67[44]*, all four DNMTs showed significantly reduced expression in EOC as compared to NO (Supplementary Fig. 9 and 10), and, more importantly, in GDHO(+) as compared to GDHO(–) EOC (Fig. 7a). In agreement, *DNMT1* and *DNMT3A* expression, when normalized to *MKI67*, inversely correlated with *LINE-1* methylation in an expanded set of EOC samples (Fig. 7b-d). In addition to DNMTs, UHRF1 is a critical component of maintenance DNA methylation[57]. Similar to *DNMTs*, after normalization to *MKI67, UHRF1* was downregulated in GDHO(+) as compared to GDHO(−) EOC (Fig. 7a). *UHRF1*, after *MKI67* normalization, also showed lower expression in EOC vs. NO (Supplementary Fig. 9e). Importantly, normalization of *DNMTs* and *UHRF1* to other established cancer proliferation markers, including *PLK1* and *BUB1[44]*, provided similar results as seen using *MKI67* normalization (Supplementary Figures 11 and 12). Together, these data suggest that GDHO(+) EOC has a reduced capacity for maintenance DNA methylation.

**Fig 7.**
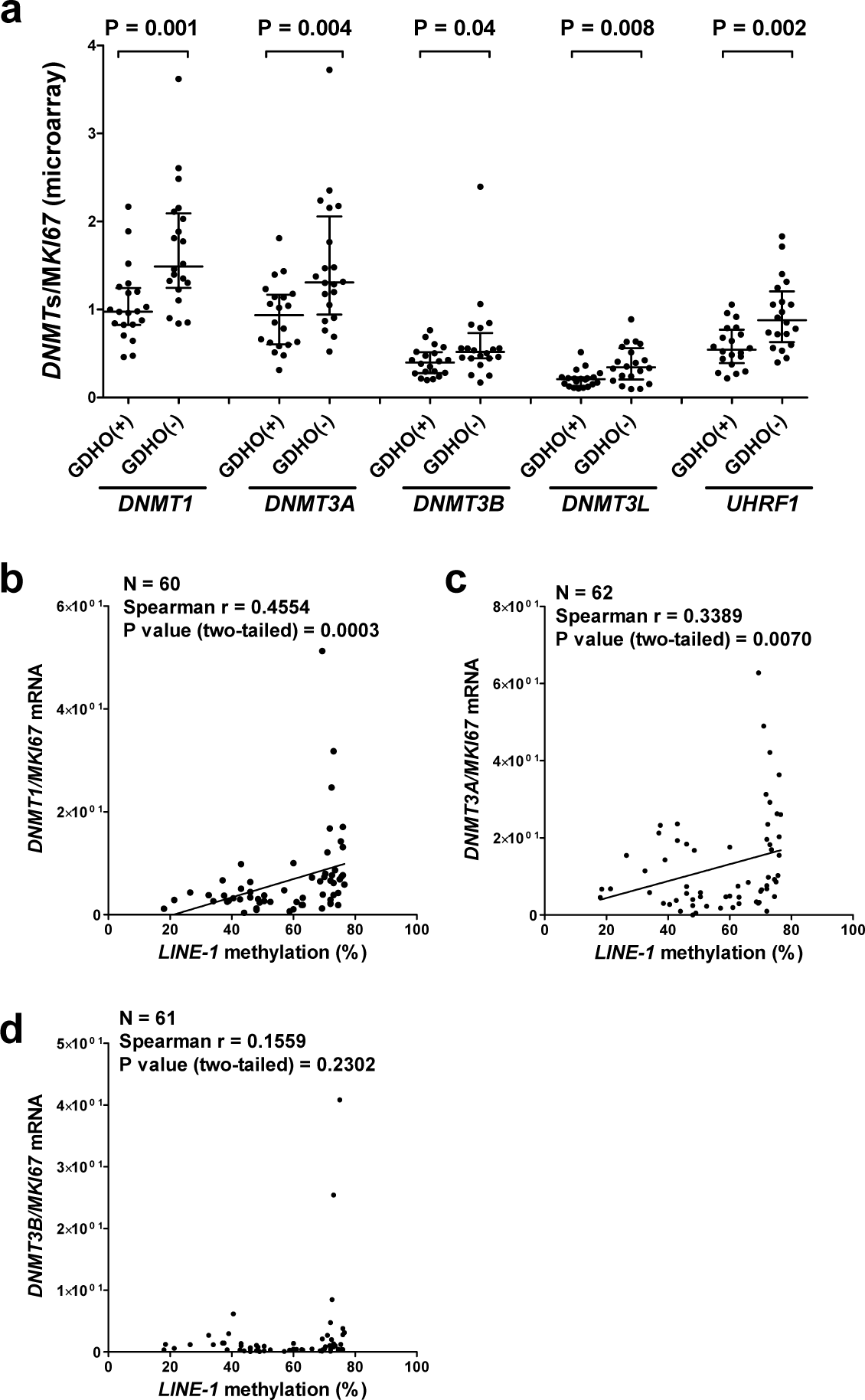
*DNMTs* and *UHRF1* expression in GDHO(+) EOC. **(a)** Affymetrix (microarray) gene expression data for *DNMT1, DNMT3A, DNMT3B, DNMT3L*, and *UHRF1*, normalized to *MKI67*, in GDHO(+) and GHDO(−) EOC. Median with interquartile range is plotted, and Mann-Whitney test p-values are shown. **(b-d)** Gene expression of **(b)** *DNMT1*, **(c)** *DNMT3A*, and **(d)** *DNMT3B*, normalized to *MKI67*, in EOC, as compared to *LINE-1* methylation in matched samples. Gene expression data were obtained by RT-qPCR, and *LINE-1* methylation was determined by bisulfite pyrosequencing. Spearman correlation analysis test results are shown.

### Repetitive element (RE) expression and hypomethylation in GDHO(+) EOC

As described above, GDHO is most commonly defined using RE methylation as a biomarker, e.g. *LINE-1* hypomethylation. As widespread RE expression has recently been reported to occur in cancer[36], we wanted to investigate the potential relationship between GDHO and RE expression. For this task, we used total RNA-seq and Methyl-seq, methods that allow precise genomic mapping of repeat sequences. Importantly, RE expression was generally elevated in GDHO(+) EOC (Fig. 8a, b). However, this effect was not uniform, but rather was class-specific (Fig. 8a; Supplementary Table 11). Most of the upregulated RE in GDHO(+) EOC were hypomethylated at their corresponding genomic loci (Fig. 8 a,b). We additionally examined the relationship between RE methylation and hypomethylated blocks. RE showed specific patterns of enrichment or depletion in hypomethylated blocks, including *LINE*, which, in most instances, was enriched in hypomethylated blocks. In contrast, *Alu/SINE* and satellite sequences were depleted from hypomethylated blocks (Table 1; Supplementary Table 12). Importantly, Methyl-seq data indicated that, as was reported previously in colon cancer[15], hypomethylation in EOC was mostly a consequence of hypomethylated block formation, rather than RE or *LINE-1* hypomethylation (Fig. 8 c,d).

**Fig 8.**
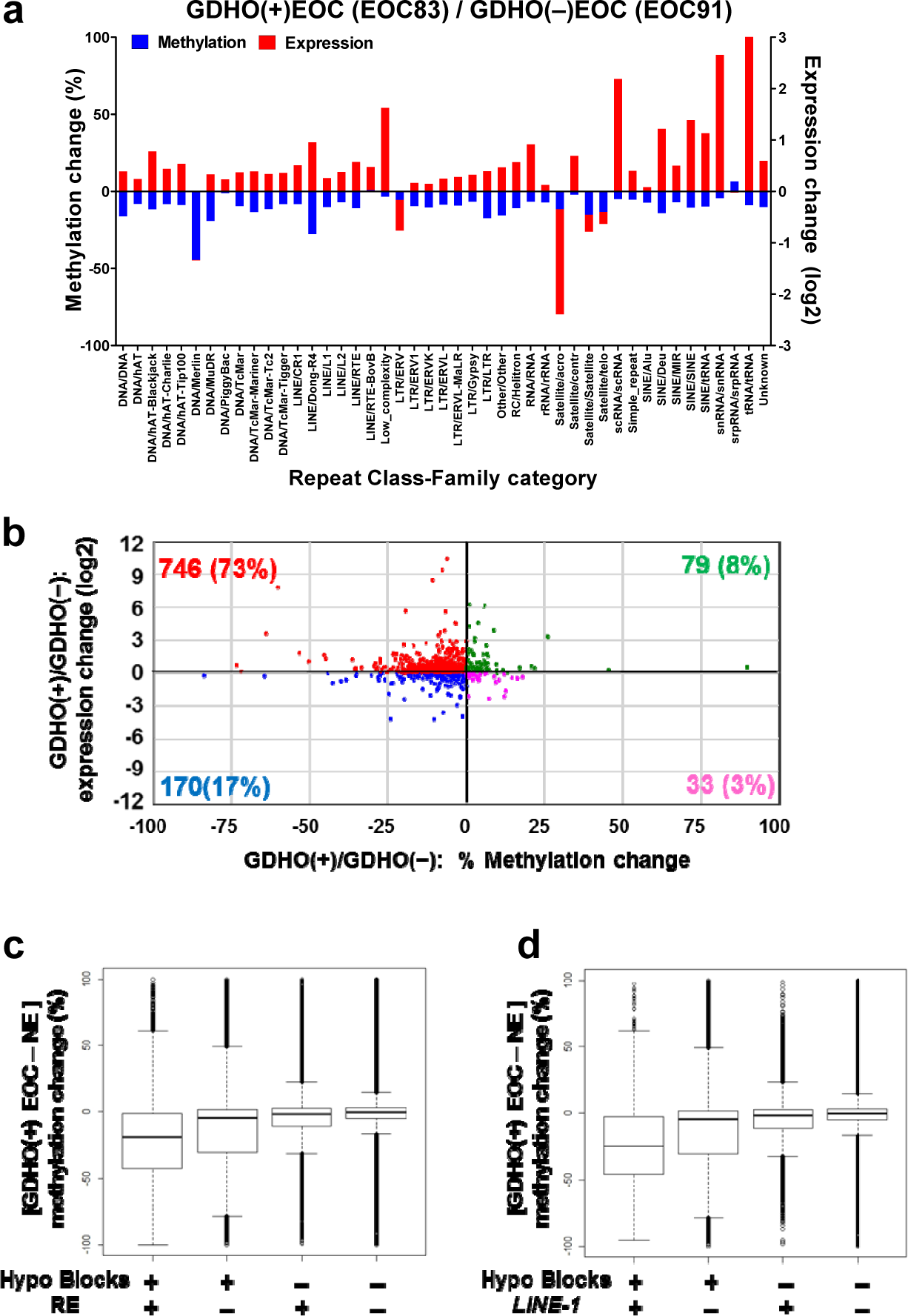
Expression and methylation of repetitive elements (RE) in GDHO(+) EOC. **(a)** Comparison of the expression and methylation of major sub-classes of REs in GDHO(+) vs. GDHO(–) EOC. Expression data were obtained from RNA-seq and methylation data from Methyl-seq. EOC83 and EOC91 are matched for stage and grade (stage 3C, grade 3). **(b)** RE expression vs. methylation in GDHO(+) vs. GDHO (–) EOC, using the same samples as in panel **a**. RE expression and methylation were determined by RNA-seq and Methyl-seq, respectively. Data points correspond to the average % methylation and log2 expression change across RE families. The results indicate that most RE families show increased expression and decreased methylation in GDHO(+) EOC. **(c)** DNA hypomethylation at hypomethylated blocks and/or all RE, in GDHO(+) as compared to NE, as determined by Methyl-seq. **(d)** DNA hypomethylation at hypomethylated blocks and/or all *LINE-1* elements, in GDHO(+) as compared to NE, as determined by Methyl-seq. In **c-d**, the median + interquartile range values are indicated. The results indicate that most hypomethylation is due to hypomethylated block formation rather than RE or *LINE-1* hypomethylation.

### Elevated chromosomal instability (CIN) and copy number alterations (CNA) in GDHO(+) EOC

To further understand the oncogenic consequences of GDHO, we focused our attention on CIN, a hallmark of EOC[38], using two complementary approaches. We first used a previously reported 25-gene expression signature of CIN to interrogate our gene expression data[58]. This signature (CIN25) was highly elevated in GDHO(+) as compared to GDHO(–) EOC, both when analyzing all samples as well as in diseased-matched samples (Fig. 9a). Second, we directly measured CNA in an additional set of 40 disease-matched GDHO(+) and GDHO(–) EOC (all HGSOC), using copy number/SNP arrays. The data revealed that CNA was significantly elevated in GDHO(+) EOC (Fig. 9b). Moreover, CNA was uniformly high in GDHO(+) EOC (with one exception), while it was highly variable in GDHO(–) EOC (Fig. 9b). These data suggest that other important contributors to CIN may be variably present in GDHO(−) EOC. Finally, we observed significant enrichment of CNA in hypomethylated blocks (Fig. 9c), suggesting that hypomethylated block formation could promote the acquisition of CIN in EOC.

**Fig 9.**
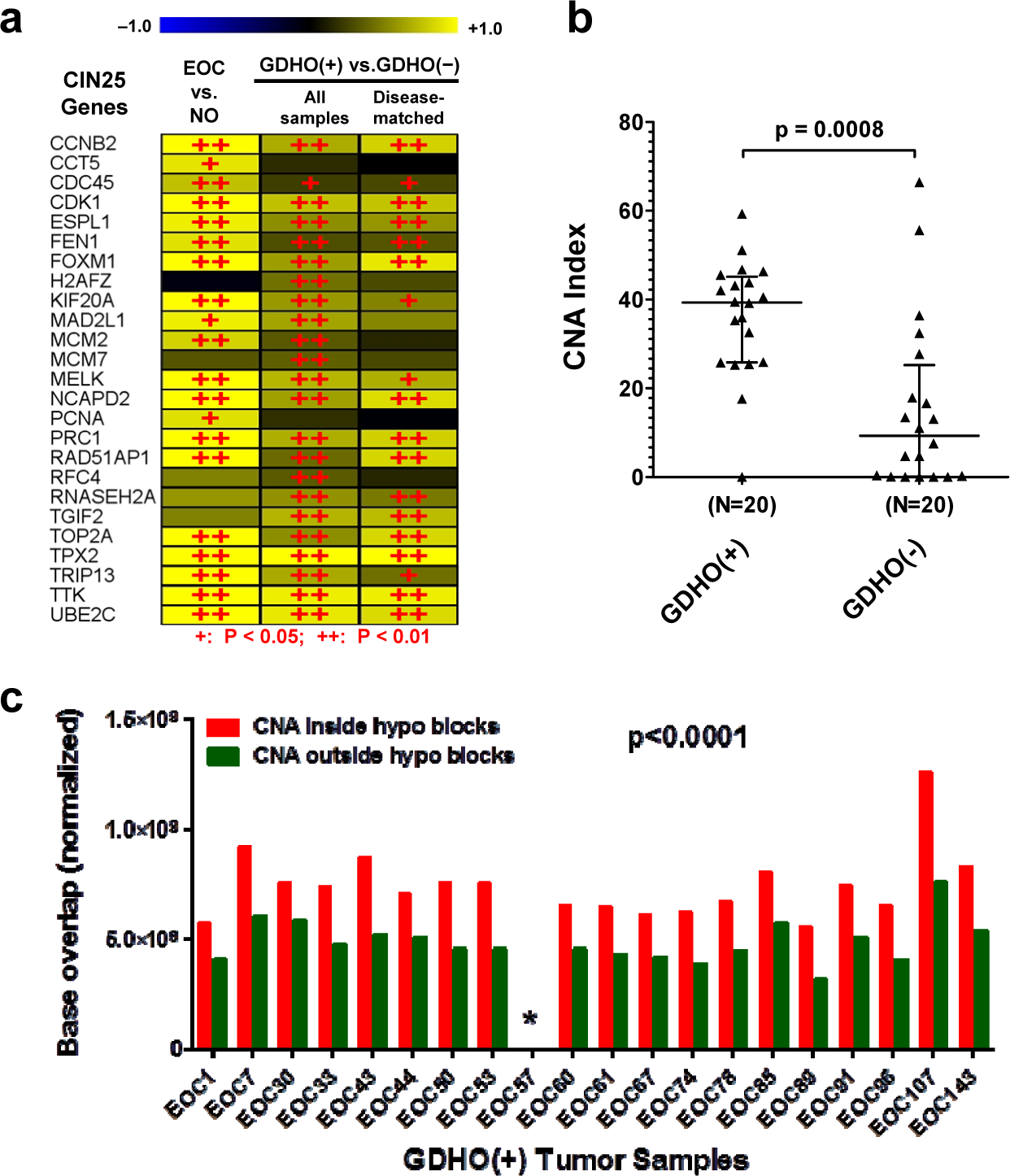
Chromosomal instability (CIN) and copy number alterations (CNA), are significantly increased in GDHO(+) EOC, and CNA are enriched at hypomethylated blocks. **(a)** Expression of CIN25 signature genes in the indicated sample comparisons. Gene expression was determined by Affymetrix microarrays, and log2 fold change values are shown. + p<0.05,++ p<0.01. **(b)** CNA index in GDHO(+) EOC vs. GDHO(–) EOC. EOC samples are disease-matched and represent HGSOC. CNA were determined using Affymetrix Cytoscan HD chips as described in *Methods*. Median with interquartile range is plotted, and the Mann Whitney p-value is shown. **(c)** CNA are enriched in hypomethylated blocks. The total base pair overlap between CNA and hypomethylated blocks, or CNA outside hypomethylated blocks (i.e. non-hypomethylated block) regions, in GDHO(+) EOC. Hypomethylated blocks were determined by Methyl-seq analysis of GDHO(+) EOC vs. normal epithelia (NE; OSE + FTE average), and CNA were determined for each tumor sample, as described in panel **b**. Hypomethylated block and non-hypomethylated block regions were normalized according to the percent of genomic coverage for each. The two-way ANOVA p-value is shown. The asterisk indicates that the values are too low to be visible on the scale.

## Discussion

We report an evaluation of the molecular pathology associated with GDHO in EOC. We used *LINE-1* hypomethylation as a biomarker for GDHO based on our earlier work [9], and observed that GDHO was associated with disease progression (advanced stage and grade) and reduced survival (disease-free and overall). By analyzing the mRNA transcriptome, RE expression, the DNA methylome, and copy number variation in parallel, we uncovered major molecular features of GDHO in EOC. These include: i) distinct patterns of gene expression, including activation of key drivers of EOC cell proliferation including *CCNE1* and *FOXM1*, ii) widespread activation of CG antigen genes; iii) deregulated expression of epigenetic regulators and histone genes, iv) increased expression of RE, often in conjunction with hypomethylation at the associated loci, and v) increased CNA and CIN. Each of these features has oncogenic potential that are likely to contribute to disease progression and poor prognosis. In addition to the characteristics described above, DNA methylome analyses revealed the formation of hypomethylated blocks, which were not ubiquitous in EOC, but rather occurred most dramatically in GDHO(+) EOC. Importantly, all known genes involved in maintenance DNA methylation displayed significantly reduced expression, relative to cell proliferation markers, in GDHO(+) EOC. Together, increased proliferation, hypomethylated block formation (which predominated at late-replicating regions), and decreased *DNMT* and *UHRF1* expression strongly suggests that passive demethylation is a key contributor to GDHO in EOC.

DNA hypomethylation was the first epigenetic alteration discovered in cancer [2]. Early seminal studies showed that DNA hypomethylation can target oncogenes, coincides with overall loss of 5mdC, and is enriched in metastatic tumors [4-6,59]. Despite its early recognition, the mechanisms underlying GDHO are only recently emerging. The recent discovery of TET-assisted oxidative DNA demethylation, involving intermediates including 5-hydroxymethylcytosine (5hmC), suggests TET activation as a potential mechanism for GDHO. However, this appears to be unlikely, as TETs are often mutated or downregulated in cancer, and cancer tissues show dramatically reduced levels of 5hmC [60]. Other possible mechanisms underlying GDHO have been proposed. For example, we earlier reported that an increased (*BORIS*) *CTCFL/CTCF* ratio correlates with *LINE-1* hypomethylation in EOC [9]. However, we found that BORIS overexpression was insufficient to promote DNA hypomethylation [61]. Other recent studies have suggested that UHRF1 overexpression (in hepatocellular carcinoma), and PIWI protein repression (in testicular cancer) may contribute to GDHO [62,63]. However, in the current study, we found no evidence for these mechanisms in EOC. In contrast, the data presented herein suggest that GDHO in EOC results, at least in part, from a proliferation-dependent process. The most statistically significant functional pathways upregulated in GDHO involved cell growth and proliferation, and, this was true for both unselected and disease-matched samples. We hypothesize that increased proliferation may overwhelm the capacity for EOC cells to perform faithful maintenance DNA methylation. Consistent with this model, a study of replication timing and DNA methylation found that late-replicating regions were generally hypomethylated and that this accumulated over many cell divisions [64]. Also supportive of this model is the recent observation of a short delay between DNA replication and the completion of maintenance methylation in cancer cells [65]. Finally, a recent landmark study demonstrated that DNA hypomethylation in cancer is concentrated at late-replicating domains that are partially methylated in normal tissues, and enriched in solo “WCGW” CpG sites [28].

A passive model for GDHO predicts that DNA hypomethylation should be concentrated at the genomic regions that are the most difficult to methylate, e.g. heterochromatic and late replicating regions, including LADs; and this is precisely what we and others have observed [14-17,27,28]. The association of GDHO with LADs should be viewed with caution, however, as LADs have not been simultaneously mapped with DNA methylation in parallel. In this context, a recent study found a link between DNA hypomethylation and late replication timing, and also showed that the association of each of these elements with LADs was secondary [66].

Because early data indicated that DNMT expression is increased in cancer, it has long been assumed that DNMTs show a gain of function in cancer, which could in turn promote the epigenetic silencing of tumor suppressor genes. However, this concept is paradoxical to the global reduction of methylation frequently observed in cancer [2]. Moreover, tumor suppressor functions for DNMTs have been demonstrated [67,68]. Because DNMTs are cell cycle regulated [56], and cancers, especially at late stages, often have an elevated proliferation index, we hypothesized that it is appropriate to normalize DNMT expression to cell proliferation, in order to more accurately gauge the functional capacity of maintenance methylation. Remarkably, after normalization to canonical cancer proliferation markers such as *MKI67*, we observed significantly reduced expression of all *DNMTs* and *UHFR1* in GDHO(+) EOC. In addition to *DNMT1*, reduced expression of *DNMT3A/3B/3L* would be expected to impair maintenance methylation, as these enzymes contribute to maintenance methylation of heterochromatic genomic regions, presumably including LOCKs (large organized chromatin K9 modifications) and LADs [69,70]. In addition to reduced expression of methylation enzymes, a recent report noted that human cancers also show uncoordinated expression of these genes as compared to normal tissues [71]. Although we did not directly measure DNMT protein expression in the current study, it was recently reported that DNMT protein expression is downregulated in high malignant potential vs. low malignant potential EOC, and, moreover, that reduced DNMT protein expression correlates with poorer survival in EOC [72]. These critical observations affirm our mRNA expression data and are also consistent with our data showing that GDHO correlates with reduced survival.

The link observed between *FOXM1* and GDHO is of interest. While FOXM1’s association with GDHO may be indirect due to its role in promoting cell cycle progression, this pathway may be particularly relevant in HGSOC [38,73,74]. A recent study reported that FOXM1 overexpression leads to DNA methylation changes in oral keratinocytes, including a global hypomethylation phenotype similar to that observed in a relevant squamous cell carcinoma cell line [46]. Coupled with our current data, an investigation of the potential mechanistic connection between FOXM1 and DNA hypomethylation in EOC is warranted.

A seminal early study demonstrated that expression of the prototype CG antigen gene *MAGEA1* was linked to global DNA hypomethylation [8]. Cadieux et al. later reported a link between GDHO, *MAGEA1* expression, and cell proliferation in glioblastoma [75]. In the current study, we measured expression of all CG genes in EOC using microarrays and found that approximately one-fifth were activated in conjunction with GDHO. The lack of ubiquitous expression of all CG genes in GDHO(+) EOC suggests additional regulatory mechanisms for CG antigen expression, e.g. oncogenic transcription factors [10]. More generally, dramatically enhanced expression of CG antigens in association with GDHO suggests that patients harboring such tumors could be an optimal group for CG antigen-directed immunotherapy. Notably, immune checkpoint modulators, such as α-CTLA4 or α-PD1/PD-L1 antibodies, require tumor antigens for their clinical activity [76]. These agents, as well as CG antigen-directed vaccines, may be most effective in patients with GDHO(+) tumors [10,45,77,78]. It will be relevant to test this concept in prospective clinical studies, in which the diagnostic tumor is used for GDHO assessment.

Pivotal work using murine models and human cell lines established the initial link between DNA hypomethylation and genomic instability [20,21]. This link has also been observed in studies using primary human tumor samples, but early studies focused on a limited number of CpG methylation sites and chromosomal alterations. More recently, large scale genomics approaches have been applied to this question and have reported clear associations of DNA methylation loss with various forms of cancer genomic instability [27,28,45]. In the current study, we comprehensively determined CNAs in HGSOC, which among human cancers studied to date, shows the most extreme level of CIN [79]. GDHO(+) tumors showed significantly and consistently higher levels of CIN, and most notably, CNAs were enriched in hypomethylated blocks. Consistent with our data, a recent study reported that chromosomal breakpoint regions in breast cancer co-localize with DMRs, which were found to be typically hypomethylated in cancer [80].

In summary, this study provides new insight into the nature of DNA methylation loss in EOC. Future work should concentrate on using this knowledge to better understand the role of epigenetic alterations in the pathogenesis of EOC, and to spur the development of therapeutic approaches to target tumors showing this defect.

## Methods

### Human tissue samples

NO, OSE, FTE, and EOC samples were obtained from patients undergoing surgical resection at Roswell Park Cancer Institute (RPCI) using Institutional Review Board Protocol I-215512. These samples have been described previously[9,43], and clinicopathological information is provided in Supplementary Table 13. NE controls (OSE and FTE) were obtained from patients without malignancy, and EOC samples contained >90% neoplastic cells. Processing of frozen tissue samples was described previously[40]. Supplementary Table 14 lists all sample groups used in this study and the genomic and epigenomic analyses conducted on each.

### DNA, RNA, and protein extractions

Genomic DNA was isolated using the Puregene Tissue Kit (Qiagen). Total RNA was purified using TRIzol (Invitrogen). Total cellular protein was extracted using RIPA buffer. Extractions were performed as described previously[9,40].

### Cell lines and drug treatments

IOSE-121 and OVCAR3 cells were described previously[81]. Cells at ∼50% confluence were treated with 1μM decitabine (DAC) (day 0), passaged at day 2, re-treated with 1μM DAC at day 3, and harvested for RNA and DNA extractions at day 5. PBS was used as the vehicle control. The data describing DAC treatment in Figure 4 is compiled from treatment of these two cell lines.

### Gene expression microarrays

Affymetrix HG 1.0ST array analysis was performed at the University at Buffalo Center of Excellence in Bioinformatics and Life Sciences. Microarray probe cell intensity data (.cel) were normalized using the Affymetrix Expression Console (version 1.3.0.187) software running the RMA workflow. We used a regularized t test analysis of control versus treatment comparisons using a Bayesian approach to estimate the within-treatment variation among replicates, using Cyber-T software. Expression heat maps were created using the TM4 microarray software suite Multi Experiment Viewer (MeV) software hierarchical clustering routine based on a Pearson correlation metric and average linkage. Functional identification of gene networks was performed using IPA (Ingenuity Systems). Gene set enrichment analysis (GSEA, http://www.broadinstitute.org/gsea/index.jsp) was used to determine the enrichment of the FOXM1 transcription factor network (http://www.broadinstitute.org/gsea/msigdb/cards/PID_FOXM1PATHWAY.html), genes involved in the G2/M checkpoint (http://www.broadinstitute.org/gsea/msigdb/cards/HALLMARK_G2M_CHECKPOINT.html), and CG genes (http://cancerimmunity.org/resources/ct-gene-database) in GDHO(+) EOC vs. GDHO(−) EOC.

### Reverse Transcriptase-Quantitative PCR (RT-qPCR)

RT-qPCR was performed as described previously[9]. Briefly, DNase-treated RNA was converted to cDNA, and samples were analyzed in triplicate using a BioRad CFX Connect system and the SYBR green method. Expression data were normalized to *18s rRNA*. All primer sequences are provided in Supplementary Table 15.

### Western blot analysis

Western blotting was performed as described previously[9], and used the rabbit polyclonal primary anti-FOXM1 antibody (K19, Santa Cruz Biotechnology, sc-500, 1:500 dilution), and the goat anti-rabbit IgG-HRP secondary antibody (Santa Cruz Biotechnology, sc-2004, 1:5000 dilution). Ponceau S (Acros) staining was used as an additional loading control.

### Bisulfite clonal sequencing and pyrosequencing

Genomic DNA was converted using the EZ DNA Methylation Kit (Zymo Research). Bisulfite sequencing was accomplished as described previously[82] and DNA sequences were analyzed using Lasergene (DNASTAR). *LINE-1* pyrosequencing was performed as described previously[40], using the PSQ HS96 System (Qiagen). All primer sequences are provided in Supplementary Table 15.

### DNA methylome analyses

Illumina Infinium 450K BeadChip analysis was performed at the RPCI Genomics Core and the University of Utah Genomics Core Facilities. Agilent SureSelect Methylome sequencing, a targeted solution hybridization bisulfite sequencing method (SHBS-seq)[55], was performed at the UNMC Epigenomics Core Facility. This method encompasses ∼15% of genomic CpG sites (3.5 × 10^6^ CpGs). High-throughput sequencing of library tags was performed at the UNMC Sequencing Core Facility, using an Illumina HiSeq 2000 Genome Analyzer. Sequence tags were aligned to the human genome (hg19) using the methylated sequence aligner Bismark (http://www.bioinformatics.babraham.ac.uk/projects/bismark/). The coverage of different genomic regions by Methyl-seq is shown in Supplementary Table 8. Differentially methylated regions (DMR) corresponded to genomic regions of any length containing ≥ 3 CpG sites, with ≥ 1 CpG site showing a mean methylation change of >20% at p ≤ 0.05. RnBeads was used to analyze both 450K and Methyl-seq data, in order to define methylation changes including DMR, promoters, CpG sites, and other genomic elements (http://rnbeads.mpi-inf.mpg.de/).

### Determination of EOC hypomethylated blocks

We defined hypomethylated blocks essentially as described previously[16]. Briefly, we used RnBeads to determine hypomethylated blocks using Methyl-seq data for NE control [(FTE + OSE average (N=2)] versus GDHO(+) EOC (N=2), using a ≤ 0.05 false discovery rate (FDR) of 5 kb tiled regions, with a ≥ 35% average methylation decrease. We combined regions that were ≤ 250kb distance apart, and selected only final blocks containing ≥ 5 CpGs.

### Correlation of EOC hypomethylated blocks with genomic features

Spatial correlations were calculated between the reference hypomethylated block genomic intervals determined from GDHO(+) EOC Methyl-Seq data and the query genomic intervals of specific genomic features, using the R package GenometriCorr (http://genometricorr.sourceforge.net/). CTCF, EZH2 and SUZ12 transcription binding sites were acquired from the UCSC Genome Browser hg19 table browser wgEncodeRegTfbsClusteredV3 table. *LINE-1, SINE-Alu*, and satellite repeat genomic locations were acquired from the UCSC Genome Browser hg19 table browser RepeatMasker. LAD genomic locations were acquired from the UCSC Genome Browser hg19 table browser NKI LADs (Tig3) database. Early and late replicating genomic regions were acquired from the UCSC Genome Browser hg19 table browser Replication Timing by Repli-chip from ENCODE/FSU table IMR90 1, wgEncodeFsuRepliChipH1hescWaveSignalRep1. Histone modifications were acquired from the ENCODE ChIP-seq of mammary epithelial cells, using H3K9me3 (accession ENCFF001SWV), H3K4me3 (accession ENCFF001SXB), H3K27ac (accession ENCFF001SWW), H3K36me3 (accession ENCFF001SWY), and H3K27me3 (accession ENCFF001SWX).

### Total RNA sequencing (RNA-seq)

RNA-seq was performed at the UNMC Sequencing Core Facility using the TruSeq Stranded Total RNA kit (Illumina) and an Illumina HiSeq 2000 Genome Analyzer. The starting material was 1.0 µg total RNA/sample. The resulting sequence tags were aligned to the UCSC Genome Browser reference human genome (hg19) mRNAs and REs using the software TopHat (v2.0.8)[83]. Cufflinks (v2.1.1) and Cuffdiff (v2.1.1) were used to estimate the expression values and determine differential expression of REs[83].

### Genomic copy number analysis

Genomic copy number analysis was performed at the UNMC Sequencing Core Facility, using Affymetrix Cytoscan HD microarrays. The total number and size (segments) of CNA per sample were determined using the Affymetrix Chromosome Analysis Suite (ChAS) Software. A CNA index was calculated for each sample based on the percent of the genome that resulted in either a copy number loss or gain. Base overlap (CNA inside hypomethylated blocks) and non-overlap (CNA outside hypomethylated blocks) between each EOC CNA segment and hypomethylated blocks was determined using the Bedtools intersect routine (http://bedtools.readthedocs.org/en/latest/content/tools/intersect.html).

### Correlation of *LINE-1* methylation with EOC patient survival

Overall survival was defined as the number of months between the diagnosis date and death, and patients still alive were censored at their date of last follow-up. For disease-free survival, patients who were alive and disease-free were censored at the date of the last visit. *LINE-1* methylation, as determined by bisulfite pyrosequencing, was segregated into three groups as shown in Fig. 1c,d, and survival was compared using Kaplan Meier analyses. The null hypothesis of no difference in the survival distributions was assessed using the Logrank test. The age-independent association of *LINE-1* methylation with survival was tested using a proportional hazards model.

### Genomic data deposit and public access

Upon publication of this paper, all data from each genomic parameter platform will be deposited into the NCBI Gene Expression Omnibus (GEO). Illumina Infinium Human Methylation 450K BeadChip array data will be deposited as *.idat files. High throughput sequencing analysis Total RNA-seq and Agilent’s SureSelect Target Enrichment Bisulfite Sequencing data will be deposited as *.fastq files. Affymetrix GeneChip Human Gene 1.0 ST and Cytoscan HD microarray data will be deposited as *.cel files.

## Data Availability

All data is provided in the contents of the manuscript.

## Non-standard abbreviations

CG gene: cancer germline or cancer testis gene
CIN: chromosomal instability
CNA: copy number alteration
DAC: decitabine, 5-aza-2’-deoxycytidine
DEG: differentially expressed gene
DE-CG gene: differentially expressed CG gene
DMC: differentially methylated CpG site
DMR: differentially methylated region
DNMT: cytosine DNA methyltransferase
EOC: epithelial ovarian cancer
FDR: false discovery rate
FTE: fallopian tube epithelia
GDHO: global DNA hypomethylation
GSEA: gene set enrichment analysis
HGSOC: high grade serous ovarian cancer
IPA: Ingenuity Pathway Analysis
LAD: lamina-associated domain
NE: normal epithelia (OSE + FTE)
NO: bulk normal ovary
OSE: ovarian surface epithelia
PCA: principal component analysis
RE: repetitive DNA elements
RMA: robust multichip average
TET: Ten-eleven translocation methylcytosine dioxygenase
TF: transcription factor

## Acknowledgements

We thank Drs. Jennifer Black and Michael Green for critical review of the manuscript. We thank the following core facilities for support: University of Buffalo Center of Excellence Genomics and Bioinformatics, RPCI Bioinformatics and Genomics, UNMC DNA Sequencing and Microarray and Bioinformatics and Systems Biology (both received support from NIGMS 8P20GM103427 and P20GM103471), and Epigenomics, and University of Utah Genomics.

## Funding

A.R.K. was supported by NIH RO1CA116674, DOD OCRP W81XWH-12-1-0456, Otis Glebe Medical Research Foundation, Betty J. and Charles D. McKinsey Ovarian Cancer Research Fund, Fred & Pamela Buffett Cancer Center (NCI Cancer Center Support Grant P30 CA036727), and RPCI Alliance Foundation. C.J.B. was supported by NIH T32CA009476. S.N.A. was supported by NIH T32CA108456. KO was supported by NCI Cancer Center Support Grant P30 CA016056, NIH 1R01CA158318, and RPCI-UPCI Ovarian Cancer SPORE P50CA159981, and the Roswell Park Alliance Foundation.

## Author Contributions

W.Z. and A.R.K. designed the research; W.Z., D.K., and C.J.B. performed the research; S.N.A., and K.O. contributed reagents; W.Z., D.K., S.P., C.G., C.J.B., A.M., and A.R.K. analyzed the data; and W.Z, D.K., and A.R.K. wrote the paper.

## Competing Financial Interests

The authors declare no competing financial interests.

## Notes

### Competing Interest Statement

The authors have declared no competing interest.

### Funding Statement

All funding is reported in the contents of the manuscript.

